# Clinical Characteristics of Recurrent-positive Coronavirus Disease 2019 after Curative Discharge: a retrospective analysis of 15 cases in Wuhan China

**DOI:** 10.1101/2020.07.02.20144873

**Authors:** Lan Chen, Zhen-Yu Zhang, Xiao-Bin Zhang, Su-Zhen Zhang, Qiu-Ying Han, Zhi-Peng Feng, Jian-Guo Fu, Xiong Xiao, Hui-Min Chen, Li-Long Liu, Xian-Li Chen, Yu-Pei Lan, De-Jin Zhong, Lan Hu, Jun-Hui Wang, Zhen-Yu Yin

## Abstract

In China, the patients with previously negative RT-PCR results again test positive during the post-discharge isolation period. We aimed to determine the clinical characteristics of these “recurrent-positive” patients. We retrospectively reviewed the data of 15 recurrent-positive patients and 107 control patients with non-recurrent, moderate COVID-19 treated in Wuhan, China. Clinical data and laboratory results were comparatively analyzed. We found that recurrent-positive patients had moderate disease. The rate of recurrent-positive disease in our hospital was 1.87%. Recurrent-positive patients were significantly younger (43(35-54) years) than control patients (60(43-69) years) (P=0.011). The early LOS (length of stay in hospital before recurrence) was significantly longer in recurrent-positive patients (36(34-45) days) than in control patients (15(7-30) days) (P =0.001). The time required for the first conversion of RT-PCR results from positive to negative was significantly longer in recurrent-positive patients (14(10-17) days) than in control patients (6(3-9) days) (P =0.011). Serum COVID-19 antibody levels were significantly lower in recurrent-positive patients than in control patients (IgM: 13.69 ± 4.38 vs. 68.10 ± 20.85 AU/mL, P = 0.015; IgG: 78.53 ± 9.30 vs. 147.85 ± 13.33 AU/mL, P < 0.0001). Recurrent-positive patients were younger than control patients. The early LOS (length of stay in hospital before recurrence) was significantly longer in recurrent-positive group than that in control group. COVID-19 IgM/IgG antibody levels were significantly lower in recurrent-positive group than those in control group, which might explain why the virus RNA RT-PCR was positive after the initial “clinical cure”(with three times of virus RNA RT-PCR negative). The virus might not be fully eliminated because of the lower IgG level and their later replicating might result in recurrent-positive virus RNA RT-PCR.

## Introduction

In December 2019, an outbreak of a novel type of pneumonia caused by severe acute respiratory syndrome coronavirus-2 (SARS-CoV-2) occurred in Wuhan, China [1]. The disease caused by this virus, which is known as coronavirus disease 2019 (COVID-19), is highly infectious and lethal. The COVID-19 epidemic was declared a “public health emergency of international concern” by the World Health Organization (WHO) on January 31 [2,3]. On March 11, the COVID-19 outbreak was upgraded to a “global pandemic” by the WHO. As of March 29, 2020, the virus has spread to 202 countries and regions, and the number of confirmed cases has reached 581,694 worldwide, including 82,345 confirmed cases and 3,306 deaths in China [4]. Many countries have launched national and regional emergency measures to cope with this explosive epidemic. At present, the epidemic has been significantly contained in China, and Wuhan has gradually lifted its lockdown measures, but the global spread has not been significantly alleviated.

Thus far, a total of 82,345 patients have been diagnosed with COVID-19 in China, of which 3,306 patients have been declared clinically cured and discharged. In China, the discharge criteria for COVID-19 patients are as follows: (1) normal body temperature for more than 3 days, (2) significant improvement of respiratory symptoms, (3) significant improvement of acute exudative lesions on lung imaging, and (4) negative results on two consecutive reverse transcription-polymerase chain reaction (RT-PCR) tests of respiratory tract samples, such as sputum and nasopharyngeal swab, with a sampling interval of at least 24 h [5]. Patients who meet these criteria are isolated and kept under observation in the community for a further 14 days after discharge. Another RT-PCR test is conducted towards the end of the observation period, and if it is negative, the isolation and observation can be stopped, and the patient is finally allowed to go home. It has been found that during the post-discharge isolation and observation period, a few patients again show positive results on RT-PCR tests. We call these patients “recurrent-positive patients,” and they must return to their designated hospital for further treatment.

The management of recurrent-positive COVID-19 patients has become a research hot spot, and several questions related to this patient population need to be urgently answered. For instance, the proportion of recurrent-positive patients, the cause of recurrent positivity, the clinical characteristics and optimal management of recurrent-positive patients, and whether these patients are infectious all need to be determined. Since March 11, 2020, 15 recurrent-positive patients have been admitted to our ward. In this study, we analyzed the clinical data and laboratory findings of these patients as compared with a control group. We aim to describe the clinical characteristics of recurrent-positive COVID-19 patients, and hope that our findings will help guide their evaluation and treatment.

## Methods

### Ethics and consent

The study protocol was approved by the ethics committee of Optics Valley Branch of Tongji Hospital, Tongji Medical College, Huazhong University of Science and Technology, and oral consent was obtained from all enrolled patients. Tongji Hospital is one of the largest teaching hospitals located at the center of the COVID-19 epidemic in Wuhan City, Hubei Province. At the government’s behest, Tongji Hospital is responsible for the management of COVID-19 patients. Our study team is from Xiamen, Fujian Province, and is providing healthcare services at Tongji Hospital during the COVID-19 epidemic. The study protocol was also approved by the ethics committee of Xiamen Zhongshan Hospital (No xmzsyyky 2020-098).

### Participants

All COVID-19 patients were diagnosed according to the medium-term guidelines issued by the WHO [6]. Based on published guidelines [7], the diagnostic criteria for COVID-19 are as follows: (1) a positive real-time RT-PCR test for COVID-19, (2) high homology with a known COVID-19 sequence on viral gene sequencing, and (3) a positive IgM antibody assay for COVID-19 and an IgG level four times the initial value. Moderate disease is defined as fever, respiratory symptoms, and imaging analysis indicative of pneumonia. All the recurrent-positive patients enrolled in this study met the four aforementioned discharge criteria [5]. Recurrent-positive COVID-19 was defined as positive results on RT-PCR testing of a nasopharyngeal swab collected during the post-discharge isolation and observation period.

We retrospectively reviewed the data of 15 patients with recurrent-positive COVID-19 as well as 107 control patients with (non-recurrent) COVID-19 who were treated in Tongji Hospita between February 10 and March 31, 2020. All clinical data of the patients were collected, including general data, hospitalization time, time required for the first, second, and third conversions of the RT-PCR results, symptoms, preexisting diseases, and changes in organ function indexes, inflammatory factors, lymphocyte count and ratio, and COVID-19 antibody levels as determined using laboratory examination.

A negative COVID-19 result was characterized by 3 consecutive RT-PCR(-) results for each patient. The time of the first RT-PCR(-) result was recorded for further analysis. The time required for the first conversion of RT-PCR results (from positive to negative) was defined as the interval between the last positive RT-PCR result and the first negative RT-PCR result. The time required for the second conversion (from negative to positive) was defined as the interval between the last negative RT-PCR test and the detection of recurrent positivity. The time required for the third conversion (from positive to negative) was defined as the interval between the detection of recurrent positivity and the next negative RT-PCR test.

### RT-PCR for COVID-19

The COVID-19 RT-PCR detection kit (S1002) and COVID-19 nucleic acid detection kit were purchased from Beijing Youkang Hengye Biotechnology Co. Ltd. After admission, the patients’ oral and nasopharyngeal swabs were collected in the ward by trained, qualified nurses every Monday, Wednesday, and Friday morning. Two swabs were collected together in a virus-preservation tube. The SARS-CoV-2 nucleic acid genome was detected in the laboratory of Tongji Hospital. The open reading frame 1ab (*ORF1ab*) gene and the core shell protein gene N fragment were detected using real-time fluorescence RT-PCR. Positive detection was determined by reference to the operating standards for the COVID-19 RT-PCR detection method for clinical laboratories [7-9]. In addition to the two gene parameters, a typical S-type amplification curve with an amplification threshold ≤ 40 was required for a positive RT-PCR test result. If any one of these two parameters was positive, re-sampling and re-examination were performed. If any one of positive result was obtained on re-examination, the sample was determined to be positive for COVID-19. If the *ORF1ab* or N gene test result was in the gray area (suspicious), re-sampling and re-examination were performed. If the result was still in the gray area after re-examination, but the amplification curve showed obvious peaks, the sample was judged to be positive; otherwise, it was considered negative.

### COVID-19 IgM/IgG antibody assays

Serum samples were tested using the COVID-19 IgM and IgG antibody detection kits (chemiluminescence method) were purchased from Shenzhen Yahuilong Biotechnology Co. Ltd. and iflash3000 automatic chemiluminescence immunoassay. The quantitative detection of IgM and IgG in serum was carried out using a two-step indirect immunoassay. They have a positive correlation with the relative luminous intensity measured by the chemiluminescence analyzer, and the test results were assessed using the calibration coefficient of the product calibrator. Negative and positive results were defined as serum IgM and IgG concentrations of <10.0 Au/mL and >10.0 Au/mL, respectively. [10,11]

### Statistical analysis

SPSS v17.0 software was used for statistical analysis. Measurement data were tested for normality. Normally distributed measurement data were expressed as mean ± standard deviation and compared between groups by using the *t*-test. Non-normally distributed measurement data were expressed as median (interquartile range), and compared between groups by using the Wilcoxon rank sum test. Count data were expressed as percentages, and the χ^2^ test was used for between-group comparisons. Taking a = 0.05 as the test standard, we deemed differences with P < 0.05 as statistically significant.

## Results

### Baseline characteristics

As of March 30, 2020, we have treated 143 patients with suspected COVID-19. Among these 143 patients, 135 patients were confirmed to have COVID-19, including 15 patients with severe/critical illness, and 120 patients with moderate disease. Among the 135 COVID-19 patients, 2 patients died; both of these patients had severe disease at the time of admission, and died within 12 h after admission. Among the 120 patients with non-severe COVID-19, 15 patients had recurrent-positive results. One of these patients was discharged from our hospital, while the other patients were discharged from other department and transferred to our department from the isolated area. Since March 11, 2020, all 15 recurrent-positive patients were successively re-admitted to our hospital. All of these patients had non-severe disease before discharge as well as after the recurrent-positive results. The 15 patients include 8 women and 7 men. As of March 30, 2020, a total of 121 COVID-19 patients have been cured in our department, including 14 recurrent-positive patients from other hospital. Among these 107 patients, 2 patients had recurrent-positive disease, yielding an incidence rate of 1.87%. One of these patients has been resident in our ward, while the other was isolated in the community (i.e., not hospitalized).

In addition to the 15 recurrent-positive patients, this study included 107 patients with moderate, non-recurrent COVID-19 as the control group. There were 48 women and 59 men in the control group; thus, the sex ratio did not significantly differ between the recurrent-positive and control groups. Patients in the recurrent-positive group were significantly younger (median, 43 years; interquartile range, 35-54years) than those in the control group (median, 60; interquartile range, 43-69 years; P=0.01). The overall course of disease was significantly longer in the recurrent-positive group (median, 36; interquartile range, 34-45 days) than in the control group (median, 15; interquartile range, 7-30 days; P=0.001). The time required for the first conversion of the RT-PCR results (from positive to negative) was significantly longer in the recurrent-positive group (median, 14; interquartile range, 10-17 days) than in the control group (median, 6 days; interquartile range, 3-9days; P =0.011). In the recurrent-positive group, the average interval between the last negative RT-PCR test and the final diagnosis of recurrent-positive disease (i.e., the second conversion of RT-PCR results) was 12 days (interquartile range, 9–13 days). In the same group, the time required for the third conversion of the RT-PCR results was 3 days (interquartile range, 3–4.25 days), which was significantly shorter than the time required for the first conversion (P <0.001). The duration of the initial hospitalization in the recurrent-positive group was 20 days (interquartile range, 15–18 days), which was significantly longer than the duration of hospitalization in the control group (median, 13; interquartile range,11-16 days; P = 0.001). The patients in the recurrent-positive group had 2–3 negative RT-PCR results (average, 2.47 ± 0.13) prior to discharge, which is in line with the Chinese discharge criteria for COVID-19 patients. Throughout the hospitalization hormone therapy was not used in the recurrent-positive group (Table 1).

**Table 1.**
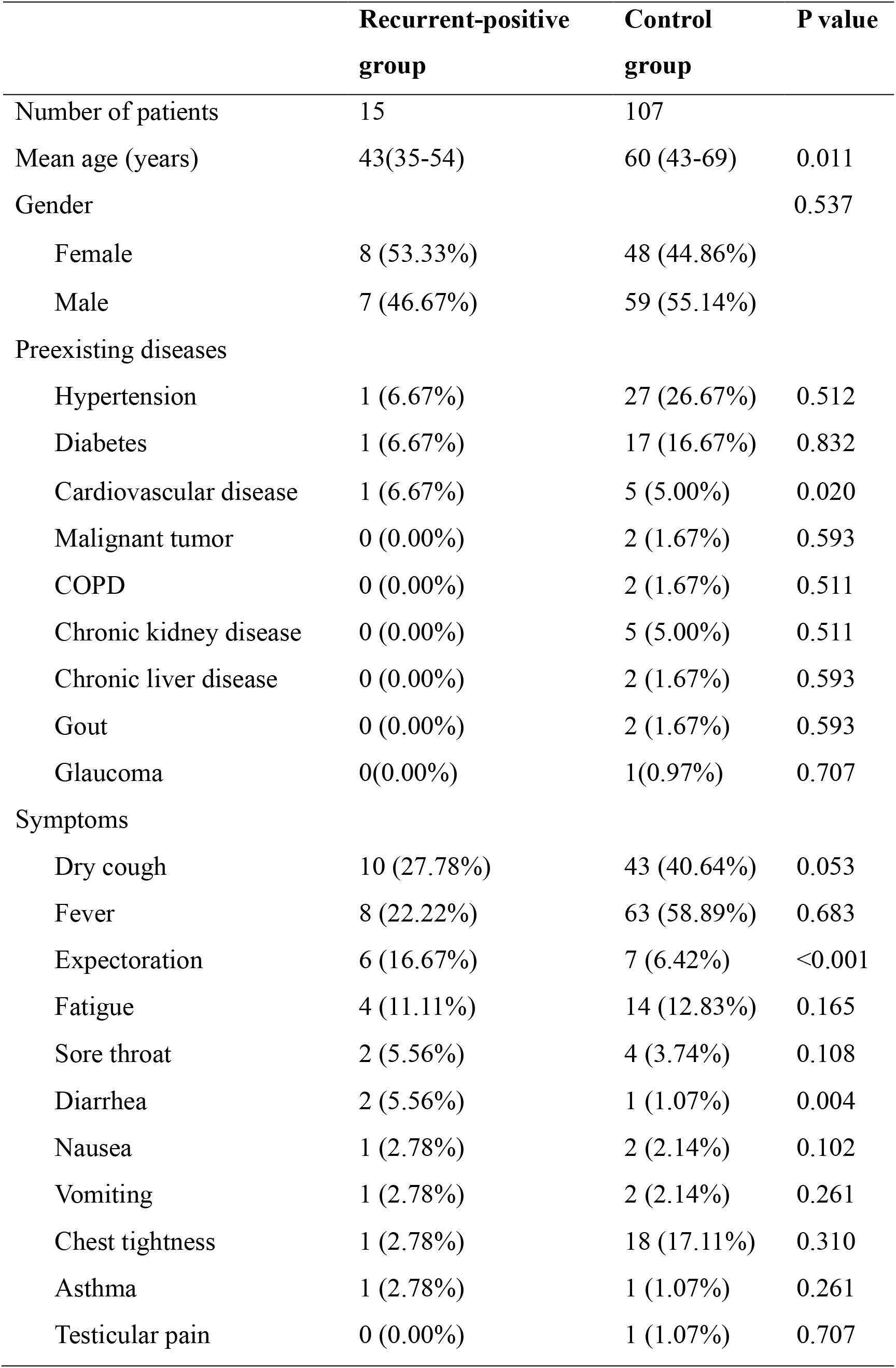
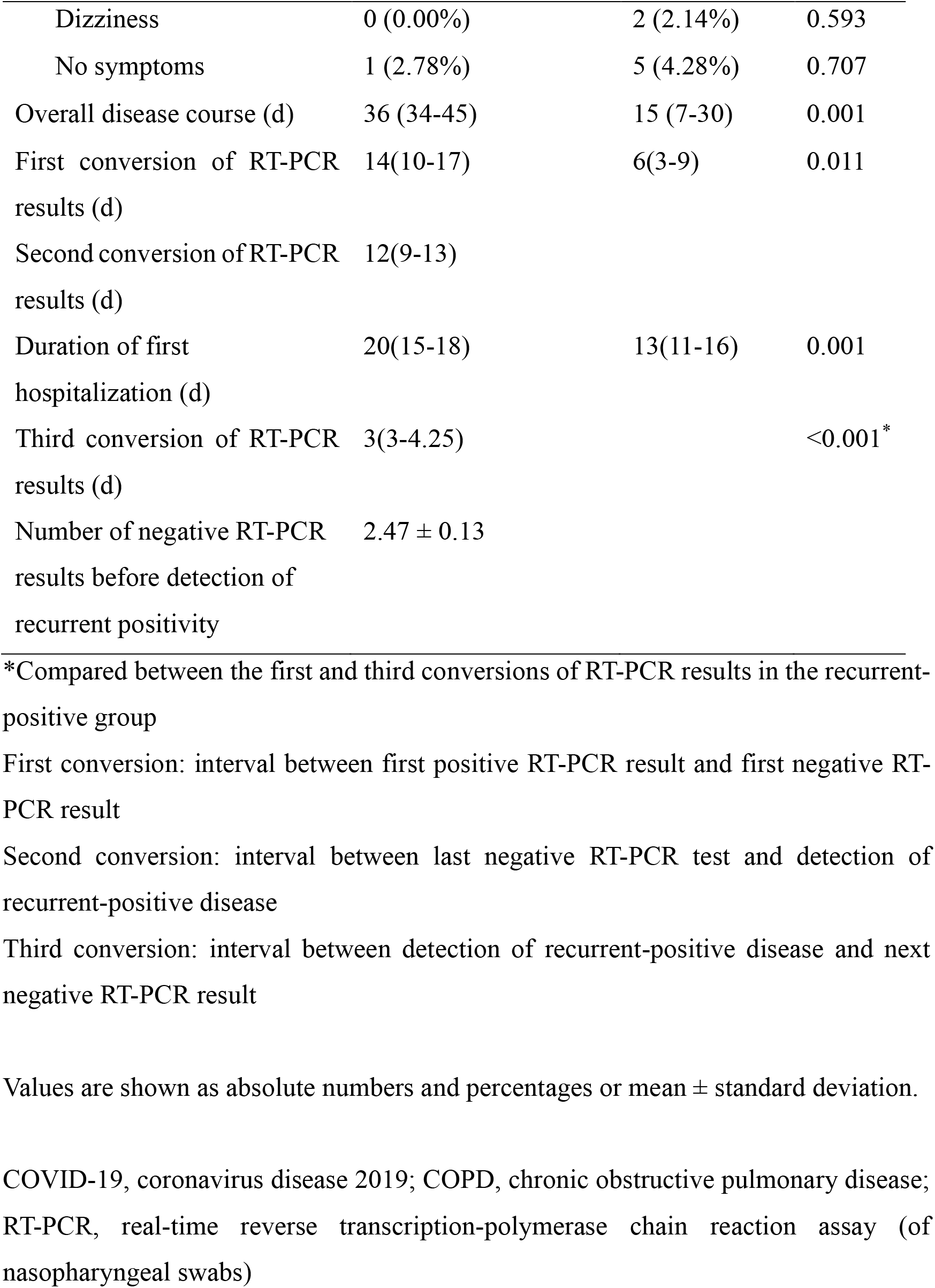
Baseline characteristics of patients with recurrent-positive and non-recurrent, moderate COVID-19

|  | **Recurrent-positive**<br>**group** | **Control**<br>**group** | **P value** |
| --- | --- | --- | --- |
| Number of patients | 15 | 107 |  |
| Mean age (years) | 43(35-54) | 60 (43-69) | 0.011 |
| Gender |  |  | 0.537 |
| Female | 8 (53.33%) | 48 (44.86%) |  |
| Male | 7 (46.67%) | 59 (55.14%) |  |
| Preexisting diseases |  |  |  |
| Hypertension | 1 (6.67%) | 27 (26.67%) | 0.512 |
| Diabetes | 1 (6.67%) | 17 (16.67%) | 0.832 |
| Cardiovascular disease | 1 (6.67%) | 5 (5.00%) | 0.020 |
| Malignant tumor | 0 (0.00%) | 2 (1.67%) | 0.593 |
| COPD | 0 (0.00%) | 2 (1.67%) | 0.511 |
| Chronic kidney disease | 0 (0.00%) | 5 (5.00%) | 0.511 |
| Chronic liver disease | 0 (0.00%) | 2 (1.67%) | 0.593 |
| Gout | 0 (0.00%) | 2 (1.67%) | 0.593 |
| Glaucoma | 0(0.00%) | 1(0.97%) | 0.707 |
| Symptoms |  |  |  |
| Dry cough | 10 (27.78%) | 43 (40.64%) | 0.053 |
| Fever | 8 (22.22%) | 63 (58.89%) | 0.683 |
| Expectoration | 6 (16.67%) | 7 (6.42%) | <0.001 |
| Fatigue | 4 (11.11%) | 14 (12.83%) | 0.165 |
| Sore throat | 2 (5.56%) | 4 (3.74%) | 0.108 |
| Diarrhea | 2 (5.56%) | 1 (1.07%) | 0.004 |
| Nausea | 1 (2.78%) | 2 (2.14%) | 0.102 |
| Vomiting | 1 (2.78%) | 2 (2.14%) | 0.261 |
| Chest tightness | 1 (2.78%) | 18 (17.11%) | 0.310 |
| Asthma | 1 (2.78%) | 1 (1.07%) | 0.261 |
| Testicular pain | 0 (0.00%) | 1 (1.07%) | 0.707 |
| Dizziness | 0 (0.00%) | 2 (2.14%) | 0.593 |
| No symptoms | 1 (2.78%) | 5 (4.28%) | 0.707 |
| Overall disease course (d) | 36 (34-45) | 15 (7-30) | 0.001 |
| First conversion of RT-PCR results (d) | 14(10-17) | 6(3-9) | 0.011 |
| Second conversion of RT-PCR results (d) | 12(9-13) |  |  |
| Duration of first hospitalization (d) | 20(15-18) | 13(11-16) | 0.001 |
| Third conversion of RT-PCR results (d) | 3(3-4.25) |  | <0.001* |
| Number of negative RT-PCR results before detection of recurrent positivity | 2.47 ± 0.13 |  |  |

\*Compared between the first and third conversions of RT-PCR results in the recurrent-positive group
First conversion: interval between first positive RT-PCR result and first negative RT-PCR result
Second conversion: interval between last negative RT-PCR test and detection of recurrent-positive disease
Third conversion: interval between detection of recurrent-positive disease and next negative RT-PCR result
Values are shown as absolute numbers and percentages or mean ± standard deviation.
COVID-19, coronavirus disease 2019; COPD, chronic obstructive pulmonary disease; RT-PCR, real-time reverse transcription-polymerase chain reaction assay (of nasopharyngeal swabs)

### Laboratory data

Lymphocyte count, hemoglobin, albumin, and glomerular-filtration rate (GFR) were all significantly higher in the recurrent-positive group than in the control group (P < 0.05). Inflammatory indexes (neutrophil percentage and C-reactive protein level) were significantly lower in the recurrent-positive group than in the control group (P < 0.05). Thus, the results of clinical evaluation were better in the recurrent-positive group than in the control group. In addition, the levels of COVID-19 IgG and IgM antibodies were significantly lower (P < 0.0001, P = 0.015, respectively) in the recurrent-positive group than in the control group (Table 2).

**Table 2.** Laboratory findings in the two groups of COVID-19 patients

|  | **Recurrent-positive**<br>**group (n = 15)** | **Control group**<br>**(n = 107)** | **P value** |
| --- | --- | --- | --- |
| SpO<sub>2</sub> (%) | 97.93 ± 0.22 | 96.89 ± 0.24 | >0.05 |
| White-cell count (*10<sup>9</sup>/L) | 6.84 ± 0.33 | 6.08 ± 0.23 | >0.05 |
| Lymphocyte count (*10<sup>9</sup>/L) | 2.16 ± 0.21 | 1.33 ± 0.06 | 0.001 |
| Neutrophils (%) | 58.12 ± 2.13 | 64.93 ± 1.32 | 0.01 |
| Platelet count (*10<sup>9</sup>/L) | 225.93 ± 10.21 | 234.43 ± 9.30 | >0.05 |
| Hemoglobin (g/L) | 140.01 ± 4.39 | 128.32 ± 1.78 | 0.023 |
| C-reactive protein (mg/L) | 2.48 ± 0.59 | 29.64 ± 4.01 | <0.0001 |
| Procalcitonin (ng/mL) | 0.06 ± 0.003 | 0.08 ± 0.01 | >0.05 |
| Albumin (g/L) | 42.23 ± 0.70 | 37.81 ± 0.60 | <0.0001 |
| Aspartate transaminase (U/L) | 35.8 ± 7.78 | 33.31 ± 1.90 | >0.05 |
| Alanine transaminase (U/L) | 24.07 ± 2.33 | 29.29 ± 2.57 | >0.05 |
| GFR (mL/min/1.73 m<sup>2</sup>) | 109.84 ± 15.05 | 101.40 ± 15.07 | 0.0002 |
| Troponin (pg/mL) | 6.41 ± 4.18 | 16.53 ± 5.84 | >0.05 |
| Myoglobin (ng/mL) | 29.76 ± 2.42 | 75.19 ± 8.91 | <0.0001 |
| CK-MB (ng/mL) | 0.63 ± 0.08 | 1.30 ± 0.18 | 0.0009 |
| D-dimer (μg/mL) | 0.44 ± 0.16 | 1.15 ± 0.27 | 0.027 |
| Interleukin-1β (pg/mL) | 3.05 ± 1.02 | 2.36 ± 0.46 | >0.05 |
| Interleukin-2R (U/mL) | 333.87 ± 33.32 | 379.82 ± 33.29 | >0.05 |
| Interleukin-6 (pg/mL) | 2.59 ± 0.70 | 5.49 ± 1.11 | >0.05 |
| Interleukin-8 (pg/mL) | 9.37 ± 1.05 | 10.51 ± 2.06 | >0.05 |
| Interleukin-10 (pg/mL) | 2.65 ± 0.73 | 2.21 ± 0.39 | >0.05 |
| TNF-α (pg/mL) | 7.75 ± 0.95 | 7.23 ± 0.48 | >0.05 |
| CD4 cell count | 877.38 ± 100.52 | 745.36 ± 45.90 | >0.05 |
| CD8 cell count | 545.00 ± 64.45 | 436.43 ± 26.37 | >0.05 |
| 2019-nCoV IgM (AU/mL) | 13.69 ± 4.38 | 68.10 ± 20.85 | 0.015 |
| 2019-nCoV IgG (AU/mL) | 78.53 ± 9.30 | 147.85 ± 13.33 | <0.0001 |

SpO<sub>2</sub>, peripheral oxygen saturation; GFR, glomerular filtration rate; CK-MB, creatine kinase-MB isoenzyme; TNF-α, tumor necrosis factor-α

### RT-PCR status and COVID-19 IgM and IgG antibody assays

The levels of COVID-19 IgG and IgM were significantly lower in the recurrent-positive group than in the control group, with the difference being especially obvious in the case of IgG. For the 14 recurrent-positive patients who were transferred from other hospitals after being discharged, no COVID-19 IgG and IgM data were available. In the one patient who was re-admitted to our hospital, the level of COVID-19 IgM did not increase after the nucleic acid result had turned negative, but rather remained low. The IgG level was also low and continued to decrease, which may be related to the recurrent-positive status (Table 3).

**Table 3.** RT-PCR status and COVID-19 IgM and IgG antibodies in one patient with recurrent-positive disease

| Date | RT-PCR | IgM (AU/mL) | IgG (AU/mL) |
| --- | --- | --- | --- |
| 2020-2-24 | (+) |  |  |
| 2020-2-28 | (-) | 1.13 | 89.26 |
| 2020-3-1 | (-) | 1.23 | 82.39 |
| 2020-3-5 | (-) |  |  |
| 2020-3-13 | (+) |  |  |
| 2020-3-15 | (-) | 0.66 | 36.28 |
| 2020-3-19 | (-) | 0.67 | 41.92 |

## Discussion

As of March 29, 2020, 75,601 patients with COVID-19 have been cured and discharged in China. Some of the patients who were cured and discharged from the hospital developed “recurrent-positive disease.” At present, there is no statistical report on the recurrent-positive rate of COVID-19. In the next stage, we need to focus on the following issues: the clinical characteristics of such patients, the causes of recurrent-positivity, the diagnosis and treatment of recurrent-positive patients, the assessment of their infectious risk, and regular follow-up. Therefore, it is of great significance to carry out epidemiological investigations and clinical analyses of recurrent-positive patients.

Our retrospective study analyzed the clinical data of patients with recurrent-positive COVID-19 as compared to patients with non-recurrent, moderate COVID-19 and found the following: (1) The rate of recurrent-positive COVID-19 in our ward is 1.87%. (2) Patients with recurrent-positive disease were significantly younger than the control patients. (3) The general clinical status (e.g., inflammatory indexes, nutrition status, and heart, kidney, and liver function) was significantly better in the recurrent-positive group than in the control group. (4) The overall course of the disease was significantly longer in the recurrent-positive group than in the control group. (5) The time required for the first conversion of the RT-PCR results (from positive to negative) was significantly longer in the recurrent-positive group than in the control group. (6) In the recurrent-positive group, the average time required for the second conversion of the RT-PCR results (from negative to positive) was 10.20 ± 1.18 days. (7) In the recurrent-positive group, the time required for the third conversion of RT-PCR results (from positive to negative; 3.33 ± 0.33 days) was significantly shorter than that required for the first conversion (13.00 ± 1.73 days). (8) The duration of the initial hospitalization in the recurrent-positive group was significantly longer than the duration of hospitalization in the control group. (9) Each patient in the recurrent-positive group had 2 to 3 negative RT-PCR results prior to discharge. (10) No hormone therapy was used during the hospitalization in the recurrent-positive group.

The laboratory diagnosis of COVID-19 relies on the detection of the virus on RT-PCR and the detection of anti-viral IgM/IgG antibodies. Because the RT-PCR test has a false-negative rate of 30%–50% [12], the latest version of China’s COVID-19 diagnosis and treatment guidelines recommend that a positive IgM antibody assay and an IgG level of four times the initial value can be used to diagnose COVID-19 [5]. This method can be used as a supplement to RT-PCR. An absence of or a decrease in COVID-19 IgM antibody and an increase in COVID-19 IgG antibody indicate that immunity to COVID-19 has been developed and that gradual recovery may successfully occur [13-15]. Since February 28, 2020, IgG and IgM antibodies against COVID-19 have been used as diagnostic biomarkers at Tongji Hospital. Our study found that the levels of these antibodies were significantly lower in the recurrent-positive group than in the control group. Possible reasons for this may be as follows: (1) Due to immune dysfunction, the amount of protective antibodies produced after viral infection may be low, especially if the decrease of the antibodies is too fast. Thus, any viral copies remaining in the body after discharge may be able to replicate extensively under the condition of insufficient antibodies, resulting in a recurrent positive test. (2) It may be that the discharge criteria are not sufficiently stringent. Due to the high false-negative rate of RT-PCR, two negative RT-PCR tests may be insufficient; the use of three or more negative tests may lower the rate of recurrent-positive disease. (3) It may be that some recurrent-positive patients have only nucleic acid fragments, and not necessarily complete virus particles (that is, they do not necessarily have infectious virus in the body) (4) Unfortunately, we were unable to determine whether recurrent-positive patients are infectious. There is no evidence for this in our ward, and there is no relevant report in the literature. So, further observation and research are needed to settle this issue.

In conclusion, our study shows that it is important to monitor the infectious status of the recurrent-positive COVID-19 patients after their initial discharge from the hospital as well as after the treatment of the recurrent illness. To reduce the rate of recurrent-positive disease, we recommend the following measures: (1) for patients with a long course of treatment, severe symptoms, repeated treatment process, or hormone treatment, three or more consecutive, negative RT-PCR results should be used as the discharge standard. (2) The detection of IgM and IgG antibodies against COVID-19 should be used to determine the required duration of hospitalization (or post-discharge isolation), and the number of RT-PCR tests should be increased for patients who show a rapid decline in antibody levels or a low antibody titer. (3) The current recommendation of strict isolation (with masks) for 14 days after discharge patients and the reassessment of RT-PCR results at the end of the isolation period should also be followed after the second discharge of recurrent-positive patients. (5) In the treatment, infusion the plasma of convalescent patients with COVID-19 additional maybe increase the concentration of antibody in vivo, which may be effective in reducing the risk of recurrent-positive. (6) Patients with low or declining IgG antibody titers and a high risk of recurrent-positive disease may show a poor response to vaccination in the future.

## Data Availability

The corresponding author Zhen-Yu Yin are responsible for all data referred to in the manuscript.

## Author contributions

Dr Chen and Prof Yin have the assess of all the research data, and are responsible for the integrity of the data and the accuracy of data analysis.

Study design: Lan Chen, Zhen-Yu Yin

Data collection: Xiao-Bin Zhang, Zhen-Yu Zhang, Su-Zhen Zhang, Qiu-Ying Han, Lan Hu, Jun-Hui Wang

Data analysis:Jian-Guo Fu, Xiong-Xiao,De-Jin Zhong

Imaging analysis of lung: Zhi-Peng Feng

Literature search: Hui-Min Chen, Xian-Li Chen, Li-Long Liu

Manuscript drafting: Yu-Pei Lan

## Acknowledgments

This study was supported by the Medical and Health Key project fund of Xiamen(3502Z20191106) and the Project of Xiamen science and Technology Bureau fund (3502Z20194016).

## References

1. PhelanAL, KatzR, Gostin LO. The Novel Coronavirus Originating in Wuhan, China: Challenges for Global Health Governance[J]. JAMA,2020. DOI: 10.1001/jama.2020.1097. [Epub ahead of print].

2. Gorbalenya, A. E et al. The species severe acute respiratory syndrome-related coronavirus: classifying 2019-nCoV and naming it SARS-CoV-2. Nature Microbiology 2020, DOI: 10.1038/s41564-020-0695-z

3. Kupferschmidt, K.; Cohen, J. Will novel virus go pandemic or be contained?. Science 2020, 367 (6478), 610–611, DOI: 10.1126/science.367.6478.610

4. Coronavirus Disease (COVID-2019) Situation Reports 1–45; World Health Organization, 2020.https://www.who.int/emergencies/diseases/novel-coronavirus-2019/situation-reports.GoogleScholar

5. Novel coronavirus pneumonia treatment plan issued by the national health and Health Committee of People’s Republic of China (Notice on trial implementation of the Seventh Edition, the National Health Service Commission No. 2020] 184) http://www.gov.cn/zhengce/zhengceku/2020-03/04/content_5486705.htm

6. World Health Organization. Clinical management of severe acute respiratory infection when novel coronavirus(nCoV) infection is suspected:interim guidance. Published January 28,2020. Accessed January 31,2020.

7. World Health Organization (WHO). Risk communication and community engagement (RCCE) readiness and response to the 2019 novel coronavirus (2019-nCoV) Interim guidance, Interim guidance v2[EB/OL]. (2020-01-26). https://www.who.int/publications-detail/risk-communication-and-community-engagement-readiness-and-initial-response-for-novel-coronaviruses-(-ncov).

8. Centers for Disease Control and Prevention(CDC).Interim Guidelines for Collecting, Handling, and Testing Clinical Specimens from Patients Under Investigation (PUIs) for 2019 Novel Coronavirus (2019-nCoV). (2020-02-02). https://www.cdc.gov/coronavirus/2019-ncov/lab/guidelines-clinical-specimens.html?CDC_AA_refVal=https%3A%2F%2Fwww.cdc.gov%2Fcoronavirus%2F2019-ncov%2Fguidelines-clinical-specimens.html.

9. Centers for Disease Control and Prevention, Respiratory Viruses Branch, Division of Viral Diseases. Real-time RT-PCR Panel for Detection 2019-Novel Coronavirus[EB/OL]. (2020-02-04). https://www.cdc.gov/coronavirus/2019-ncov/downloads/rt-pcr-panel-for-detection-instructions.pdf.

10. PR. Hsuehab, LM. Huang, PJ. Chen, et al. Chronological evolution of IgM, IgA, IgG and neutralisation antibodies after infection with SARS-associated coronavirus. Clinical Microbiology and Infection, 2004;10(12):1062–1066. https://doi.org/10.1111/j.1469-0691.2004.01009.x

11. WZ Xu, J Li, XY He, et al. Novel coronavirus 2019 novel coronavirus IgM and IgG antibodies in the diagnosis of new coronavirus infection [J/OL]. Chinese Journal of laboratory medicine.2020,43 (2020-02-27).http://rs.yiigle.com/yufabiao/1182736.htm. DOI: 10.3760/cma.j.cn114452-20200223-00109.

12. QX Zhang, QS Zhao. High temperature inactivation process before extraction of viral nucleic acid significantly reduces the amount of detectable viral nucleic acid template. ChinaXiv, 2020.http://www.chinaxiv.org/abs/202002.00034

13. Wan SX, Yi, QJ, Fan SB, et al. Characteristics of lymphocyte subsets and cytokines in peripheral blood of 123 hospitalized patients with 2019 novel coronavirus pneumonia (NCP). doi: https://doi.org/10.1101/2020.02.10.20021832

14. PC. Y. Woo, SK. P. Lau, BH. L. Wong, et al. Longitudinal Profile of Immunoglobulin G (IgG), IgM, and IgA Antibodies against the Severe Acute Respiratory Syndrome (SARS) Coronavirus Nucleocapsid Protein in Patients with Pneumonia Due to the SARS Coronavirus. Clinical and Diagnostic Laboratory Immunology. July 2004, p. 665–668. DOI: 10.1128/CDLI.11.4.665–668.2004

15. P Li, ZY Li, SL Zhao, et al. Preliminary study on novel coronavirus pneumonia diagnosed by serum 2019-nCoV IgM and IgG antibodies [J/OL]. Chinese Journal of laboratory medicine.2020,43 (2020-03-08).http://rs.yiigle.com/yufabiao/1184360.htm. DOI: 10.3760/cma.j.cn114452-20200302-00155.

